# Non-Targeted Effects and Space Radiation Risks for Astronauts on Multiple International Space Station and Lunar Missions

**DOI:** 10.1101/2023.05.26.23290464

**Authors:** Francis A. Cucinotta

**Affiliations:** Univerity of Nevada Las Vegas, Las Vegas, NV 89154, USA

**Keywords:** space radiation, non-targeted effects, radiation cancer risk, radiation circulatory disease risks, heavy ions, galactic cosmic rays

## Abstract

Future space travel to the earth’s moon or the planet Mars will likely lead to the selection of experienced International Space Station (ISS) or lunar crew persons for subsequent lunar or mars missions. The major risk for space travel is the galactic cosmic rays (GCR) risks of cancer, circulatory diseases and detriments in cognition. However large uncertainties in risk prediction occur due to the quantitative and qualitative differences in heavy ion microscopic energy deposition leading to differences in biological effects compared to low LET radiation. In addition, there are sparse radiobiology data and absence of epidemiology data for heavy ions and other high LET radiation. Non-targeted effects (NTEs) are found in radiobiology studies to increase the biological effectiveness of high let radiation at low dose for cancer related endpoints. In this paper the most recent version of the NASA Space Cancer Risk model (NSCR-2022) is used to predict mission risks while considering NTEs in solid cancer risk predictions. I discuss predictions of space radiation risks of cancer and circulatory disease mortality for US Whites and US Asian-Pacific Islander (API) populations for 6-month ISS, 80-day lunar missions, and combined ISS-lunar mission. Results predict NTE increase cancer risks by about ∼2.3 fold over a model that ignores NTEs. US API are predicted to have a lower cancer risks of about 30% compared to US Whites. Cancer risks are slightly less than additive for multiple missions, which is due to the decease of risk with age of exposure and the increased competition with background risks as radiation risks increase. The inclusion of circulatory risks increases mortality estimates about 25% and 37% for females and males, respectively in the model ignoring NTEs, and 20% and 30% when NTEs are assumed to modify solid cancer risk. The predictions made here for combined ISS and lunar missions suggest risks are within risk limit recommendations by the NCRP for such missions.

## 1. Introduction

The International Space Station (ISS) has been continuously occupied since the year 2000 by crew sizes of 3 to 7 astronauts whom typically enjoy 6-month missions. In the past astronaut flight careers have transcended more than one space program, including astronauts from the Mercury and Gemini programs on Apollo and Skylab missions, Space Transportation System (STS) (space shuttle) program astronauts participating on NASA-MIR program and ISS missions, and an astronaut participating in Gemini, Apollo, and STS missions. The large number of ISS veterans are likely candidates for lunar and possibly Mars or other deep space missions in the future. The high occupational radiation exposures for long duration missions presents the challenge of estimating the cumulative risk, including uncertainties in the predictions over an astronaut’s career of several long duration missions.

Space radiation risk assessments carry large uncertainties due to lack of knowledge on the radiobiology of high LET radiation such as heavy ions and secondary neutrons [1–3]. The major concern for space travel is galactic cosmic rays (GCR) risks of cancer, circulatory diseases and detriments in cognition. Solar particle events are also a concern however are readily mitigated by passive shielding [4], while shielding of GCR is difficult due to their high energies. High LET radiation produces both quantitative and qualitative differences in microscopic energy deposition leading to differences in biological effects compared to low LET radiation. In addition, there are sparse radiobiology data and the absence of epidemiology data for heavy ions and other high LET radiation such as secondary neutrons and low energy protons and helium ions. Non-targeted effects (NTEs) are found in low dose radiobiology studies to increase biological effectiveness for low doses of high LET radiation [5–10] for surrogate endpoints of cancer risk. Non-targeted effects include bystander effects and changes to the tissue micro-environment, including altered biochemical signaling and the induction of chronic inflammation.

In conventional radiation protection on Earth radiation weighting factors or LET dependent quality factors (QF) are used to estimate an effective dose [11,12]. However, this approach is severely limited for higher levels of exposure and risks for long duration (>3 months) space missions, which are well above a recommended effective dose limit of 50 mSv per year [13]. The large uncertainties in radiation quality effects for heavy ions and high LET radiation suggest uncertainties in risk estimates should be evaluated and reported in radiation protection programs [1,11,14–17]. The NSCR model [15,18–20] uses track structure concepts to form a radiation quality function with parameters fit to available proton, fission neutron, helium and heavy ion data for tumor induction in animals and surrogate endpoints in cell culture models related to cancer risk. NSCR evaluates the overall uncertainty in risk models using Monte-Carlo propagation of uncertainties to combined uncertainties in epidemiology data, organ doses and fluence spectra, and dose-rate and radiation quality effects. The NSCR model was extended to include the impact of NTE’s on radiation quality descriptors in recent work [21] and in this paper, we further investigate NTE role on risk predictions.

In this paper I apply the most recent version of NSCR, denoted NSCR-2022 [21–24] to report on risk predictions of cancer and circulatory diseases for average populations of different ages that would potentially participate in multiple long duration space missions, essentially ISS and lunar missions. Risk predictions vary with the background risks for the model population and here we make predictions for representative highest (Whites) and lowest (Asian-Pacific Islanders (API)) risk groups amongst US racial and ethnic groups based on a recent assessment [23]. Because NTE’s are shown to have a dose response that deviates from linearity, we investigate the possibility of deviation from linearity on a combined exposure using 35-year old persons at first mission and 40-year old at second mission in our study.

### 2. Cancer Risk Evaluation

The instantaneous cancer incidence or mortality rates, λ_I_ and λ_M_, respectively, are modeled as functions of the tissue averaged absorbed dose *D_T_*, or dose-rate *D_Tr_*, sex, age at exposure *a_E_*, and attained age *a* or latency *L,* which is the time after exposure *L=a-a_E_*. The λ_I_ (or λ_M_) is a sum over rates for each tissue that contributes to cancer risk, λ_IT_ (or λ_MT_). These dependencies vary for each cancer type that could be increased by radiation exposure. The total risk of exposure induced cancer (REIC) is calculated by folding the instantaneous radiation cancer incidence-rate with the probability of surviving to time *t,* which is given by the survival function *S_0_(t)* for the background population times the probability for radiation cancer death at previous time, summing over one or more space mission exposures, and then integrating over the remainder of a lifetime [17,20]:

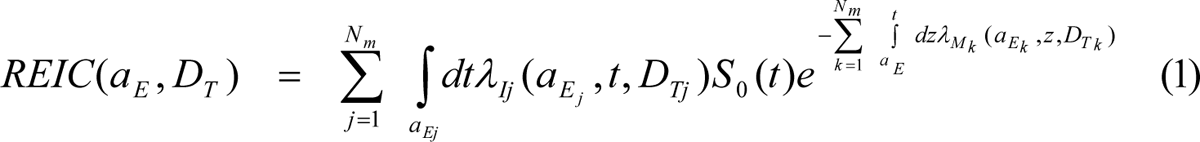

where z is the dummy integration variable. In Eq. (1), N_m_ is the number of missions (exposures), and for each exposure, *j*, there is a minimum latency of 5-years for solid cancers, 2-years for leukemia, and 5-years for circulatory diseases assumed. The upper limit of Eq. (1) is set at 100 year in the present calculations. Tissue specific REIC estimates are similar to Eq. (1) using the single term from λ_I_ of interest. The equation for risk of exposure induced death (REID) estimates is similar to Eq. (1) with the incidence rate replaced by the mortality rate (defined below) in the outer integral of Eq. (1).

The tissue-specific cancer incidence rate for an organ absorbed dose, *D_T_*, in multiplicative transfer models, denoted after adjustment for radiation quality and low dose and dose-rates through introduction of a scaling factor *R_QF_* [17,20]

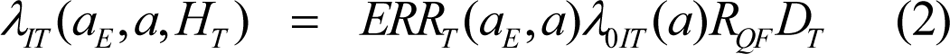

where *λ_0IT_* is the tissue-specific cancer incidence rate in the reference population, and where *ERR_T_* and is the tissue specific excess relative risk per Sievert, respectively. In previous reports we also considered a mixture model using a weighted average of the multiplicative and additive risk models. The choice of the multiplicative model in the present paper is discussed below. The tissue specific rates for cancer mortality *λ_MT_* are modeled following the BEIR VII report [25] whereby the incidence rate of equation (2) is scaled by the age, sex, and tissue specific ratio of rates for mortality to incidence in the population under study:

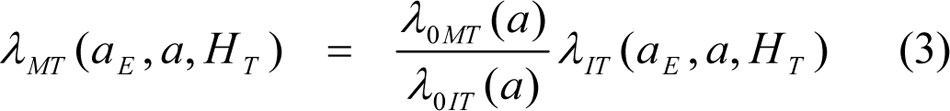

Risks predictions for circulatory disease mortality were made in the same manner as a previous report [26], however using the recent meta-analysis of ERR for mortality after low LET radiation reported by Little et al. [27,28]. This approach uses ERR from mortality data after low LET radiation exposure. Circulatory disease incidence data following radiation exposure are sparse and preclude considerations herein. There is an uncertainty related to the dependence on possible treatments reducing circulatory disease mortality with time period, host country and other factors. Such factors are likely distinct for astronauts in recent or future time periods compared to the various historical epidemiology data considered by Little et al. [28]. A dose-rate reduction effectiveness factor is not applied because the meta-analysis results used are based on low dose (<0.5 Sv) or chronic exposures. For circulatory disease risks because a relative biological effectiveness (RBE) factor is distinct from the quality factor (QF), organ dose equivalents are estimated by the NCRP for deterministic effects (tissue reactions) [11,29].

### 3. Space Radiation Quality Descriptor

The *R_QF_* is estimated from RBE’s determined from low dose and dose-rate particle data relative to acute γ-ray exposures for doses of about 0.5–3 Gy, which we denote as *RBE_γAcute_* to distinguish from *RBE_max_*, which is based on less accurate initial slope estimates for γ-rays. The basic approach is illustrated in **Figure 1**, which shows a stochastic track for an ^56^Fe ion with energy of 1 GeV/u [30]. The track is described as consisting of a so-called track core of ultra-high ionizations within 100 nm of the ions path, and a penumbra of ionizations by single energetic δ-rays extending many microns from the ions path. The quality function, *R_QF_*, considers these two contributions using a parametric approach motivated by the Katz track structure model [31,32]. However, allowing the penumbra term to follow the dose response estimated from γ-ray epidemiology studies, which is generally assumed to be linear over the relevant dose range for space missions. In addition, since we are scaling directly to the acute γ-ray responses, the dose-rate modifier is assumed to influenced by results from experimental models used in RBE determinations. The dose-rate modifier (assumed to be similar to estimate of a dose and dose-rate reduction effectiveness factor (DDREF)) adjusts the penumbra like but not the core like term of ultra-high ionizations. I note here that for chronic exposures in space it is unlikely that large regions of a genome or even individual cells would be traversed by two ion cores for time-periods of less than 1 month [33].

**Figure 1.**
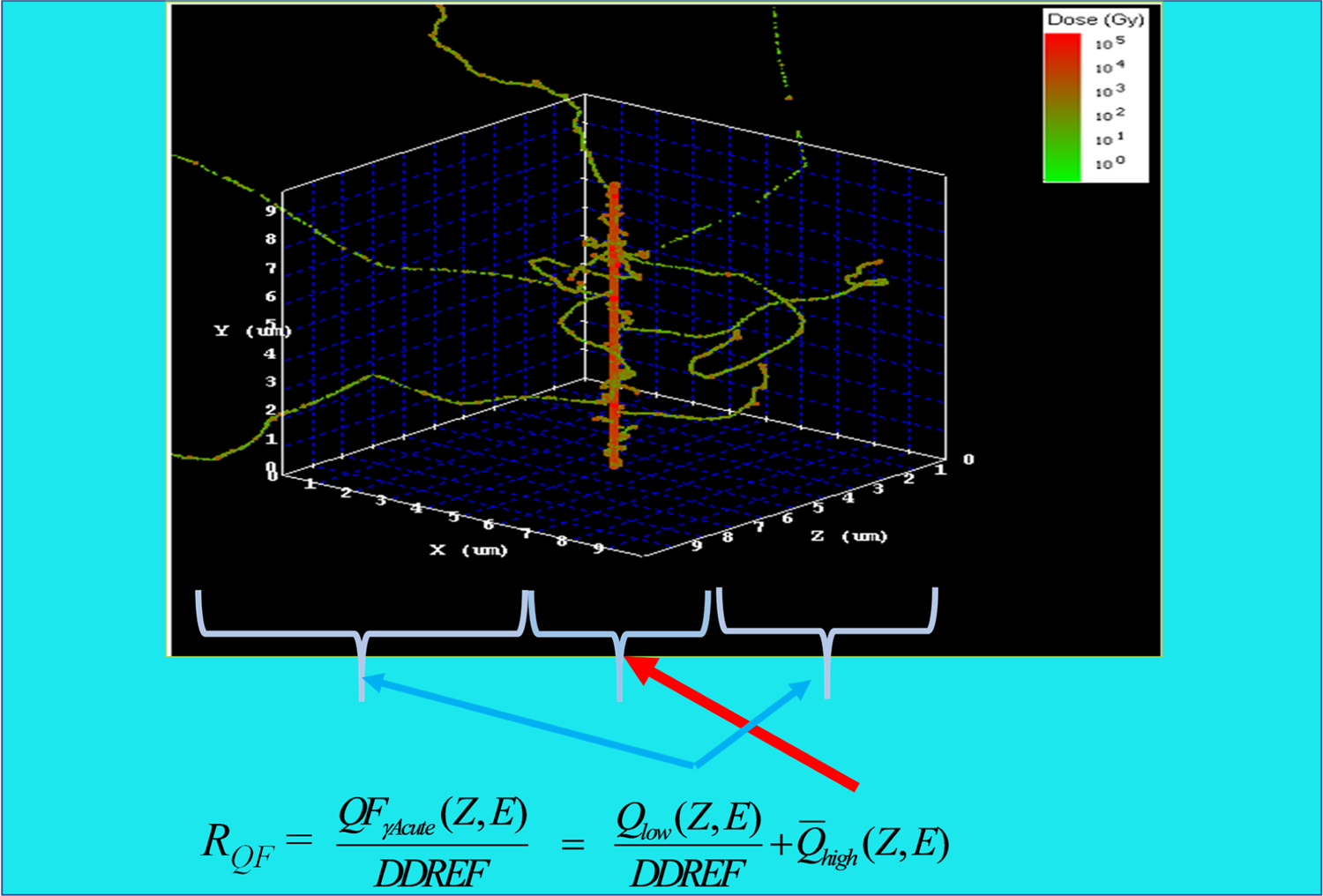
Stochastic model of ion track structure for ^56^Fe (1000 MeV/u) illustrating the NSCR-2022 model of radiation quality factor function using a separation of core and penumbra like term contributions. The penumbra term is reduced by a dose-rate modifier, while the core term with its high ionization density is assumed to be independent of dose-rate.

In this approach the scaling factor includes terms representing the penumbra-like and core-like of low and high ionization densities, respectively based on the following functions:

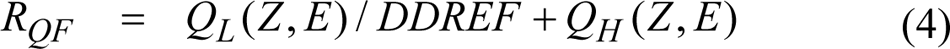

where

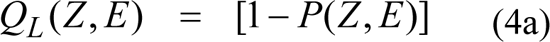

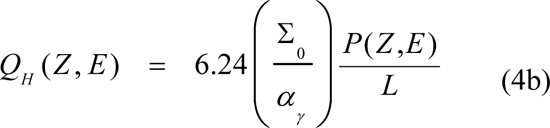

with the parametric function [20]

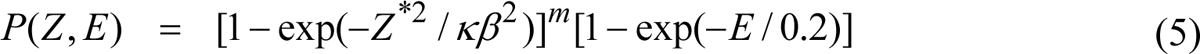

where *E* is the particles kinetic energy per nucleon, *L* is the LET in keV/μm, *Z* is the particles charge number, *Z\** the effective charge number, and *β* the particles speed relative the speed of light. The LET for protons and helium in tissue are calculated using the National Institute of Standards [34] data. Effective charge is used to scale LET of heavy ions to protons [35].

The functional form of Eq. (5) reflects predictions from both radial dose and stochastic Monte-Carlo track structure descriptions of biological action cross sections dependence on ion charge and kinetic energy [36,37], which supports its usage in the parametric NSCR model to represent available experimental data. An ancillary condition is used to correlate the values of the parameter κ as a function of *m* as [20]

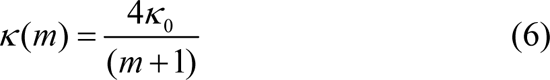

where κ_0_ is value for the most likely value *m*=3. This constraint fixes the peak effectiveness with kinetic energy for each heavy ion charge group in the model to be consistent with results from experiments [15]. In Monte-Carlo uncertainty analysis conditional sampling is used where *m* is selected from a cumulative distribution function (CDF) followed by selection of *κ(m)*, which is then distributed with a normal distribution with standard deviation (SD) shown in **Table 1**.

**Table 1.**
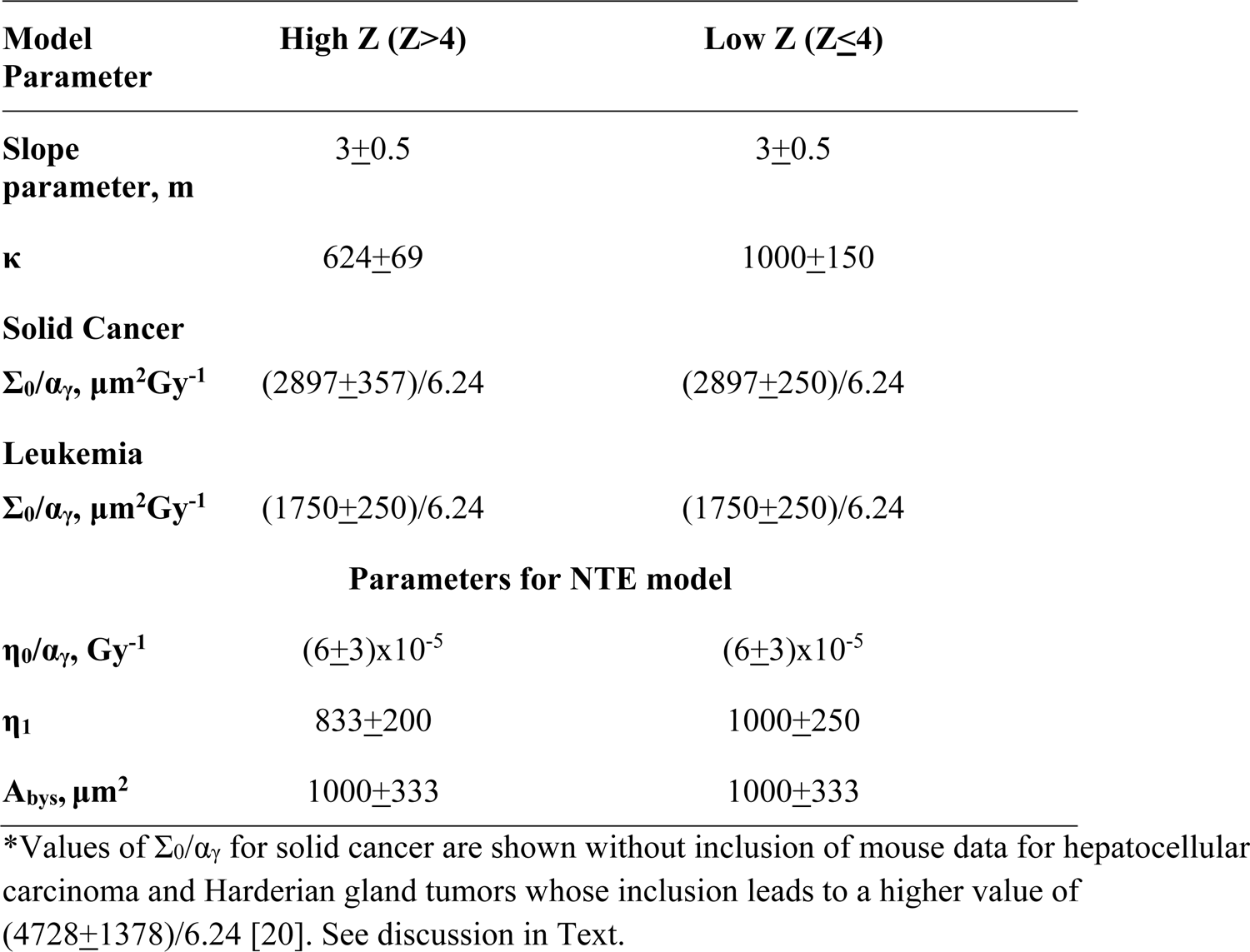
Space Radiation Quality Factor Model Parameters with standard deviation (SD) in parameter estimate*.

| Model Parameter | High Z ($Z > 4$) | Low Z ($Z \leq 4$) |
| --- | --- | --- |
| Slope parameter, m | $3 \pm 0.5$ | $3 \pm 0.5$ |
| $\kappa$ | $624 \pm 69$ | $1000 \pm 150$ |
| **Solid Cancer** |  |  |
| $\Sigma_0/\alpha_\gamma, \mu\text{m}^2\text{Gy}^{-1}$ | $(2897 \pm 357)/6.24$ | $(2897 \pm 250)/6.24$ |
| **Leukemia** |  |  |
| $\Sigma_0/\alpha_\gamma, \mu\text{m}^2\text{Gy}^{-1}$ | $(1750 \pm 250)/6.24$ | $(1750 \pm 250)/6.24$ |
| **Parameters for NTE model** |  |  |
| $\eta_0/\alpha_\gamma, \text{Gy}^{-1}$ | $(6 \pm 3) \times 10^{-5}$ | $(6 \pm 3) \times 10^{-5}$ |
| $\eta_1$ | $833 \pm 200$ | $1000 \pm 250$ |
| $A_{\text{bys}}, \mu\text{m}^2$ | $1000 \pm 333$ | $1000 \pm 333$ |

\*Values of $\Sigma_0/\alpha_\gamma$ for solid cancer are shown without inclusion of mouse data for hepatocellular carcinoma and Harderian gland tumors whose inclusion leads to a higher value of $(4728 \pm 1378)/6.24$ [20]. See discussion in Text.

The three model parameters (Σ_0_/α_γ_, κ and *m*) in Eq. (4) are fit to radiobiology data for tumors in mice or surrogate cancer endpoints as described previously [20,38]. Distinct parameters are used for estimating solid cancer and leukemia risks based on estimates of smaller RBEs for acute myeloid leukemia and thymic lymphoma in mice compared to those found for solid cancers [20]. In addition, it is noted that RBE’s for male hepatocellular carcinoma in mice are much larger than for female mice and for most other solid cancers for both male and female mice [20].

The space radiation quality model corresponds to a pseudo-biological action cross section in units of µm^2^ given by,

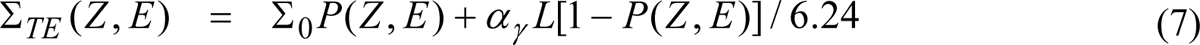

**Figure 2** compares RQF from Eq. (4) for solid cancers to the ICRP LET dependent quality factor divided by DDREF=2 as a function of kinetic energy for the major GCR components of protons, ^16^O, ^28^Si and ^56^Fe. The most important differences are at the lowest energies where protons are shown to be more effective and heavy ions less effective in NSCR-2022 compared to ICRP QF [12], and at the highest energies for heavy ions where the large penumbra of δ-rays contribution to microscopy energy deposition is predicted to reduce effectiveness, which is not described in the ICRP function. The ICRP function does not represent the branching of RBE with charge number for ions of identical LET and instead uses LET alone. In addition, the ICRP provides no uncertainty assessment for the QF, while uncertainty assessment is a main focus of NSCR model.

**Figure 2.**
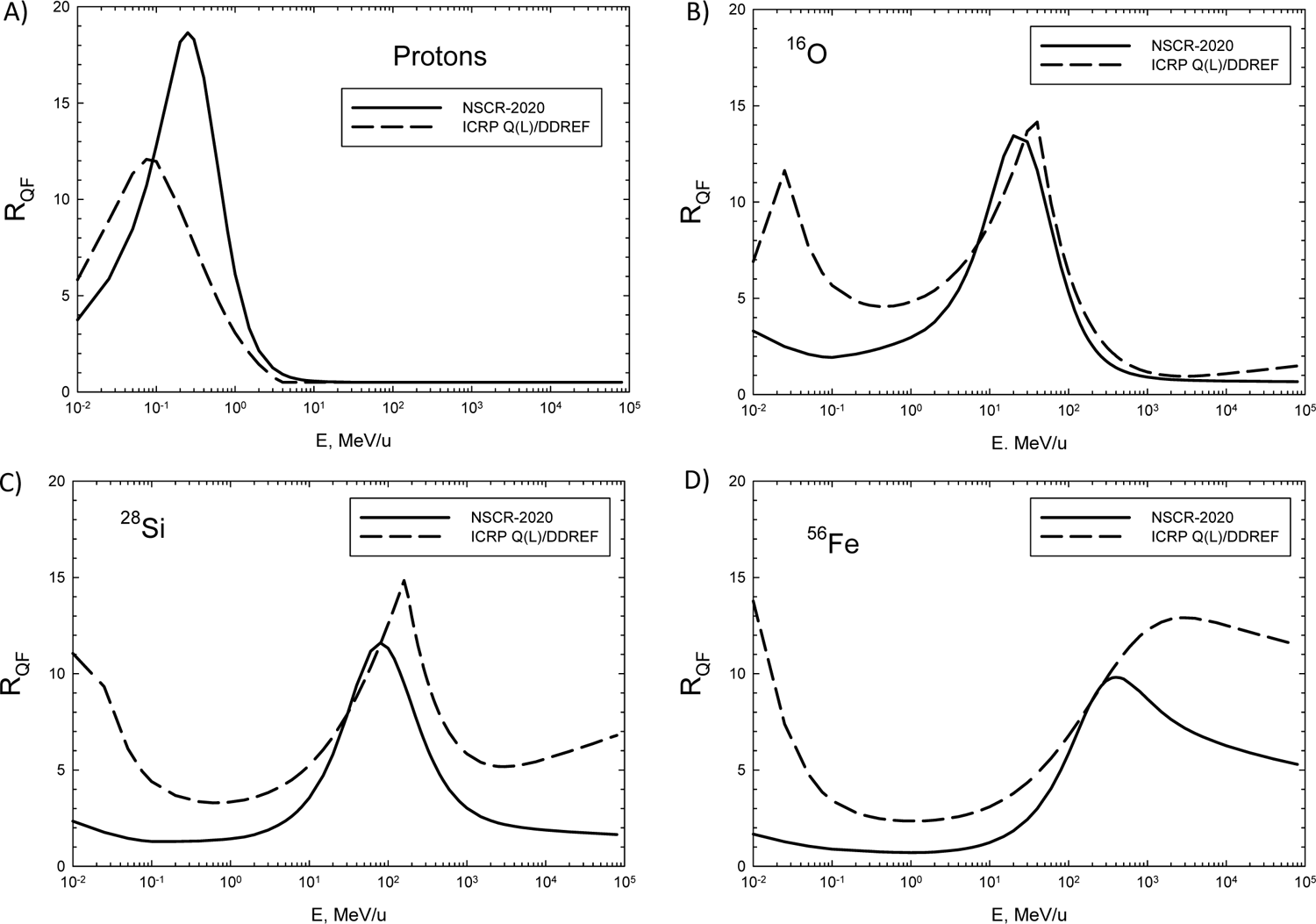
Comparison of *R_QF_* function from Eq. (4) in NSCR-2022 to ICRP Q(L) relationship divided by DDREF=2 versus ion kinetic energy in MeV/u. Panel A) Protons, B) ^16^O, C) ^28^Si and D) ^56^Fe.

### 4. Non-Targeted Effects Quality Factor Model

In the NTE model we assume the targeted effects (TE) contribution is valid with a linear response to the lowest dose or fluence considered, while an additional NTE contribution occurs [21].

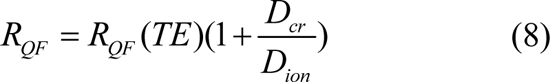

where *D_cr_* is found as the dose (or fluence) where the NTE and TE are equivalent that is given by

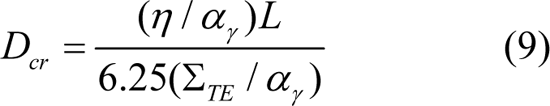

This corresponds to a pseudo-biological action cross section is given by,

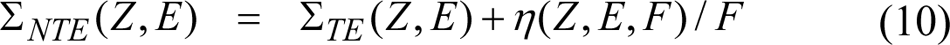

where *F* is the particle fluence (in units of 1/µm^2^) and the η function represents the NTE contribution, which is parameterized as a function of *x=Z^*2^/β^2^*as:

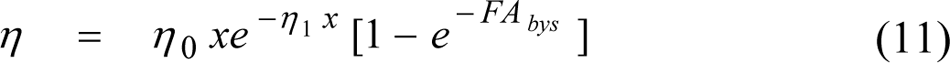

In Eq. (11) the area, *A_bys_*, reflects the region of bystander cells surrounding a cell traversed directly by a particle that receives an oncogenic signal. The parameters η_0_/α_γ_ and η_1_ are estimated from low dose radiobiology experiments for mouse Harderian gland tumor induction [8,39] and chromosomal aberrations [9]. The second factor on the right-hand side of Eq. (11) describes the “turning on” of NTE at very low doses.

The parameter *A_bys_* is difficult to estimate from reported heavy ion experiments because of lack of low dose responses (<0.01 Gy) and it is value is likely correlated with estimates of η_0_. However, several experiments with α-particles provide data to make estimates [40,41]. **Figure 3** (panels A, C, and E) shows results of fitting the model equations to experiments with α-particle induced chromosomal and chromatid aberration in wild-type CHO cells [41], and neoplastic transformation of C3H10T1/2 [40]. In **Figure 3** (panels B, D, and F) these same data are shown with the y-axis divided by the mean number of ions that traversed the cell versus the number of ions that traversed. The mean number of ions traversed is estimated as the ion fluence times the cross-sectional area of the cell nucleus (i.e. *H=FxA_cell_*), which is 62.7 and 250 μm^2^ for CHO and C3H10T1/2 cells, respectively. Using this scaling a linear response would appear as a horizontal line. There is a clear increase above linearity as the number of ion traversals decreases below unity that is reflective of a bystander effect. The magnitude of the increase is impacted by the control values found in the experiment. Here we note a null result is found for the controls and the lowest dose tested (0.0017 Gy) for chromosomal aberrations [41], while a non-zero result is found for chromatid breaks [41] and the transformation experiment [40]. The parameter *A_bys_* in Eq. (11) is estimated to vary between 900 for CHO cells and 1600 μm^2^ for the larger C3H10T1/2 cells. We note that larger distances for bystander effects are suggested in experiments using a micro-beam to target specific areas within or outside of cell nuclei [42].

**Figure 3.**
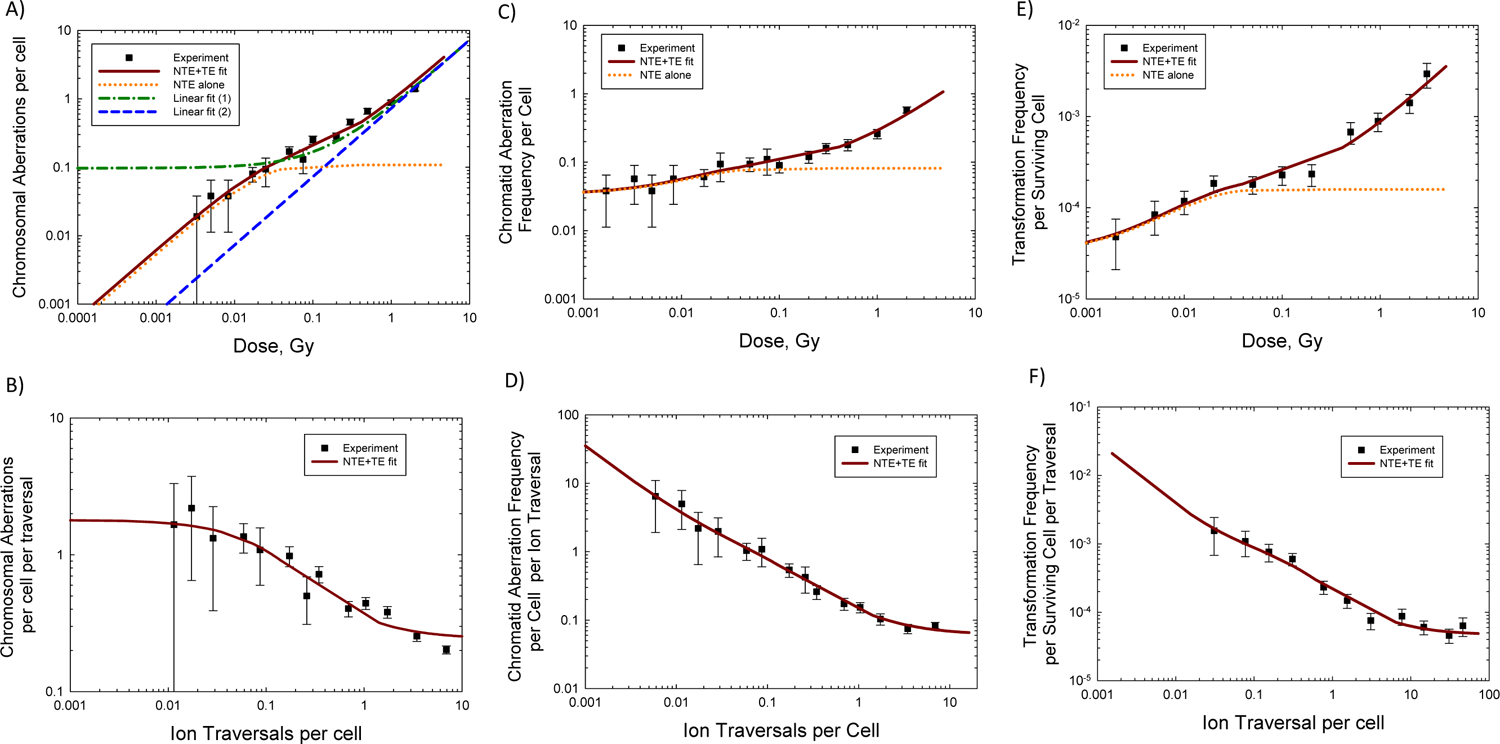
Study of dose response for chromosomal and chromatid breaks in CHO cells irradiated with α-particles (LET=112 keV/μm) (experimental data [41]), and neoplastic transformation of C3H10T1/2 cells irradiated with α-particles (LET=101 keV/μm) (experimental data [42]). Panel A) for chromosomal aberrations in CHO cells shows NTE model fits, linear fit-1 constrained to a null frequency at Dose = 0 Gy, and linear fit-2 with an unconstrained fit at Dose = 0 GY, Y=A + B Dose. Panel B) shows the data of Panel A) scaled to the mean number of ion traversals per cell versus the number of ion traversals which is equal to Fluence x A_cell_ with A_cell_ = 62 μm^2^. Panels C) and D) show results for chromatid aberrations in CHO cells, and Panels E) and F) for C3H10T1/2 cells with A_cell_=250 μm^2^.

In **Figure 4** we show a similar scaled response for Harderian gland tumors where the cell area is reported as 24 μm^2^ indicating a nuclear radius of 2.76 μm. A similar pattern is found for tumor induction as a function of heavy ion traversals per cell as that for the α-particle endpoints in **Figure 3** whereby a rapid increase occurs as the number of ion traversals decreases below unity. In **Figure 4** we also show the experimental data with background subtracted, (P-P_0_)/Hits which suggest the increasing ratio (P/H) with decreasing number of traversals is not due to the approach towards a non-zero background. For the tumor prevalence the impact of cell survival is considered, however over the low dose range (<0.2 Gy) increases in target cell survival would only increase P/H by about 20% based on cell survival experiments with heavy ions [43]. Other possibilities include radiation induced senescence or modulation of DNA damage responses reducing cancer susceptibility at higher doses to a larger extent than at lower doses. These observations which suggest bystander or other NTEs are not surprising as very few cells will survive a direct traversal by a heavy ion with LET > 100 keV/ μm of a cell nucleus. This is illustrated in **Figure 6**, which shows amorphous track model [44,45] and **Figure 7** for stochastic radiation track structures of heavy ions of high LET [30]. In Figure 7 the ^56^Fe nuclei at LET=450 keV/μm is similar to the track of the Nb beam used in the Alpen et al. experiments [39]. Clearly a large portion of a cell nucleus receives an extremely large number of ionizations for direct cell nucleus traversals of the very high LET ions (>200 keV/μm) used in the Harderian Gland experiments of Alpen et al. [39] leaving very few surviving cells. In addition, the dose-response for Harderian gland tumor induction by γ-rays does not support a high tumor prevalence from cells traversed from δ-rays produced by passing heavy ions when the ion does not traverse the cell nucleus.

**Figure 4.**
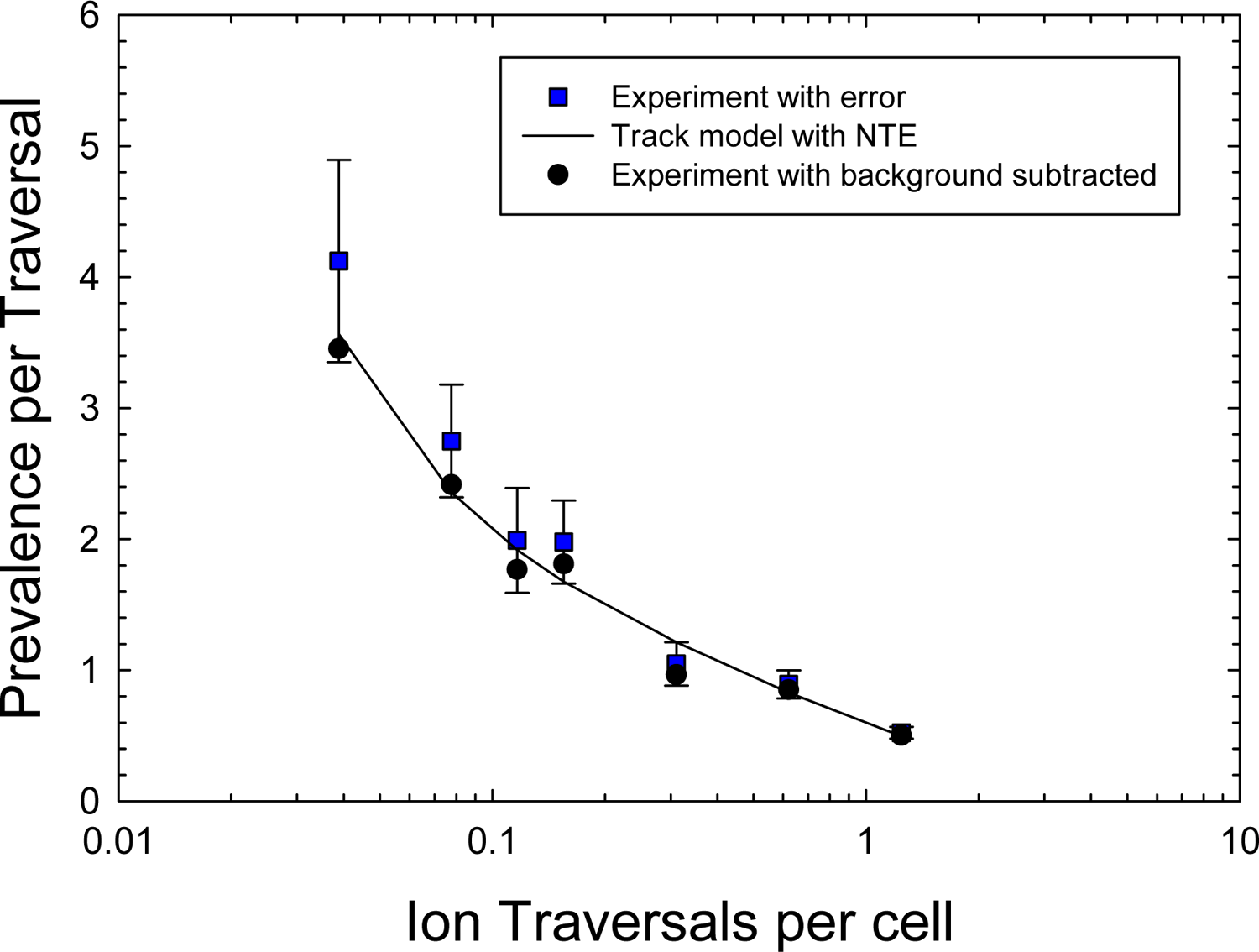
Prevalence of Harderian Gland tumors at 600 days in female B6CF_1_ mice [39] divided by mean number of ion traversals per cell nucleus versus mean number of ion traversals. Irradiation is 600 MeV/u ^56^Fe nuclei (LET=193 keV/μm). Model fit from [21]. Mean number of ion traversal is Fluence x A_cell_ with A_cell_ = 24 μm^2^.

### 5. Space Radiation Exposures Organ Exposures

GCR exposures include primary and secondary H, He and HZE particles, and secondary neutrons, mesons, electrons, and γ-rays produced over a wide energy range. We used the HZE particle transport computer code (HZETRN) [46] with quantum fragmentation model nuclear interaction cross sections [47] and GCR environmental model [15] to estimate particle energy spectra for particle type *j*, *φ_j_(Z,E)* as described previously [15,20]. For the radiation quality model described above, a mixed-field pseudo-action cross section is formed by weighting the particle flux spectra, *φ_j_(E*) for particle species, *j*, contributing to GCR exposure evaluated with the HZETRN code with the pseudo-biological action cross section for mono-energetic particles and summing over all particles and kinetic energies:

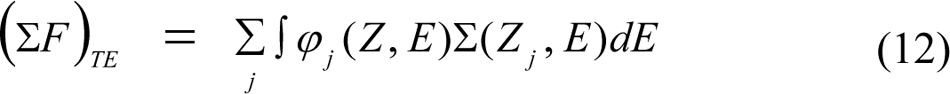

Equations for the mixed-field pseudo-action cross section in the NTE model as folded with particle specific energy spectra as [21]:

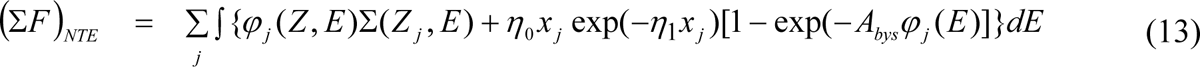

### 6. Parameter Values and Uncertainty Analysis

For the various parameters that enter into the model equations for REIC and REID, probability distribution functions (PDFs) are formulated based on experimental data to represent plausible ranges of values. The physics models for dose and dose equivalent agree with spaceflight data in low Earth orbit [13,15], in transit to Mars [48] and on the Mars surface [49] to within 15% in most comparisons as illustrated in **Figure 5**. Similar good agreement was shown for comparisons organ doses to a phantom torso experiment flown on an STS mission [13,15]. Albedo particles on the lunar or Mars surface are estimated to increase the organ dose equivalents by 10 to 20% largely due to neutrons and protons, and we use a mean increase of 15% [49]. The physics uncertainties are estimated using a normal distribution with standard deviations of 0.35 for Z≤4 ions and 0.25 for Z>4 ions. Larger deviations may occur for specific energy values of fluence spectra, however are constrained by overall uncertainty in integral quantities such as dose and dose equivalent.

**Figure 5.**
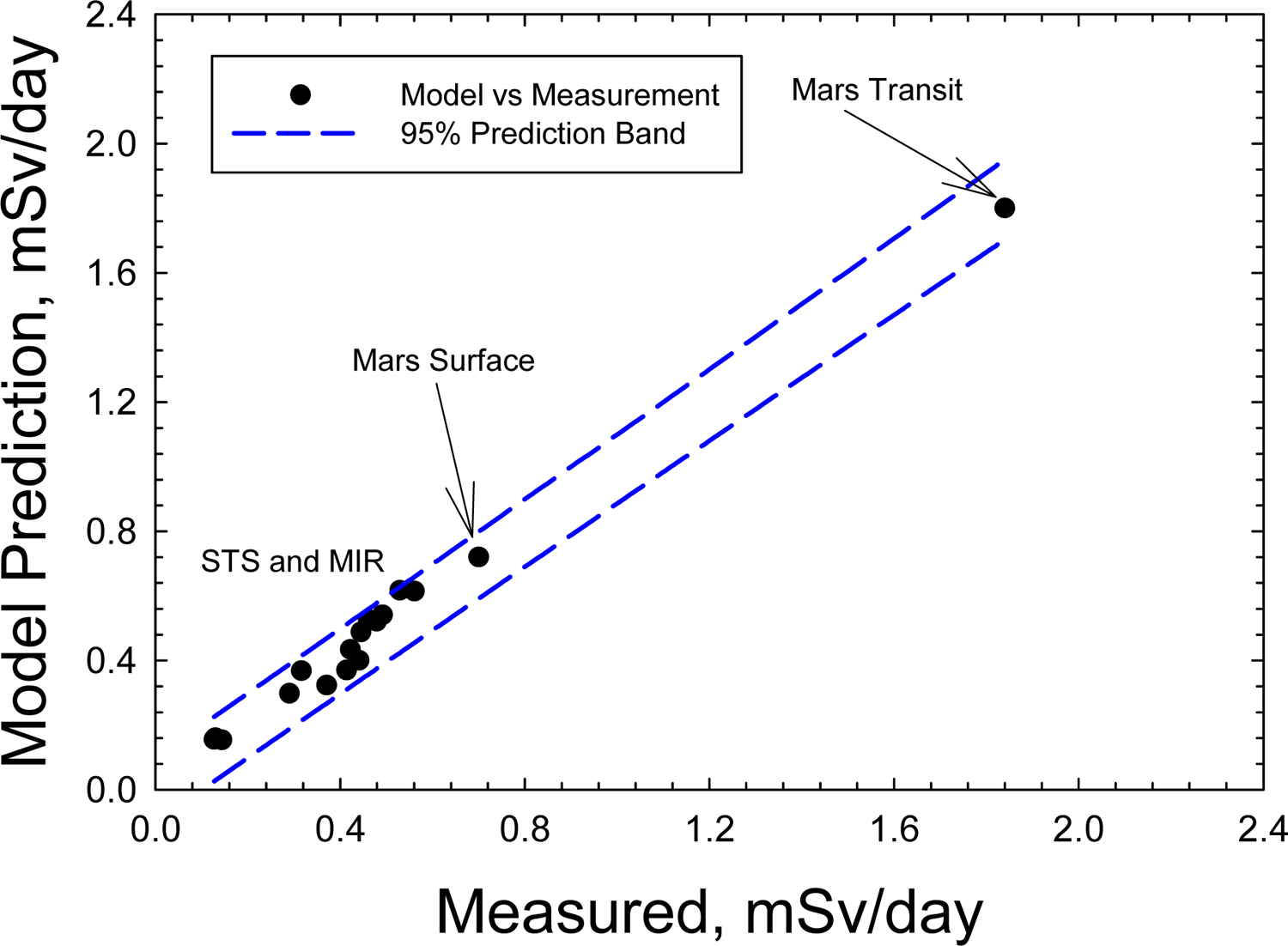
Dose equivalent from various space measurements [13,15,46,47] compared to estimates from HZETRN code used in NSCR-2022. Prediction band from SigmaPlot based on combined prediction and measurement data.

**Figure 6.**
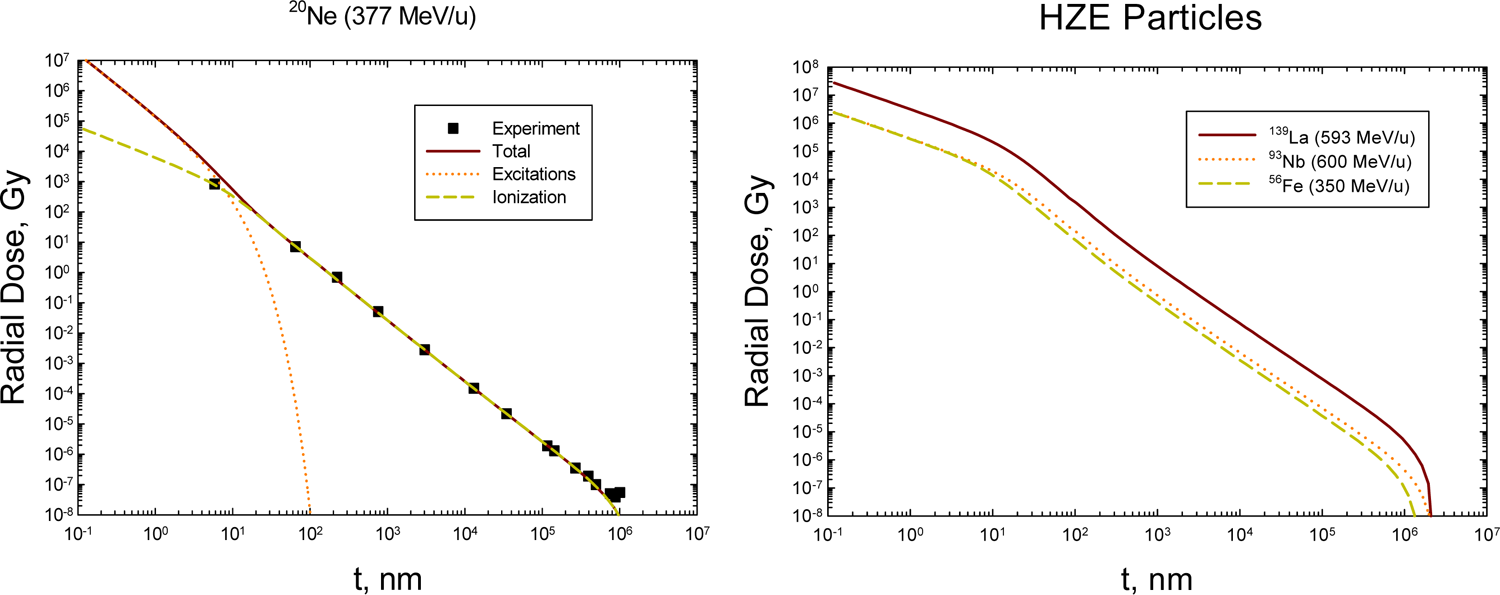
Physical descriptions of heavy ion track structure in the radial dose model of Katz [44] compared to experiments [45] in panel A) and for several ions used in the Harderian Gland experiment in panel B).

**Figure 7.**
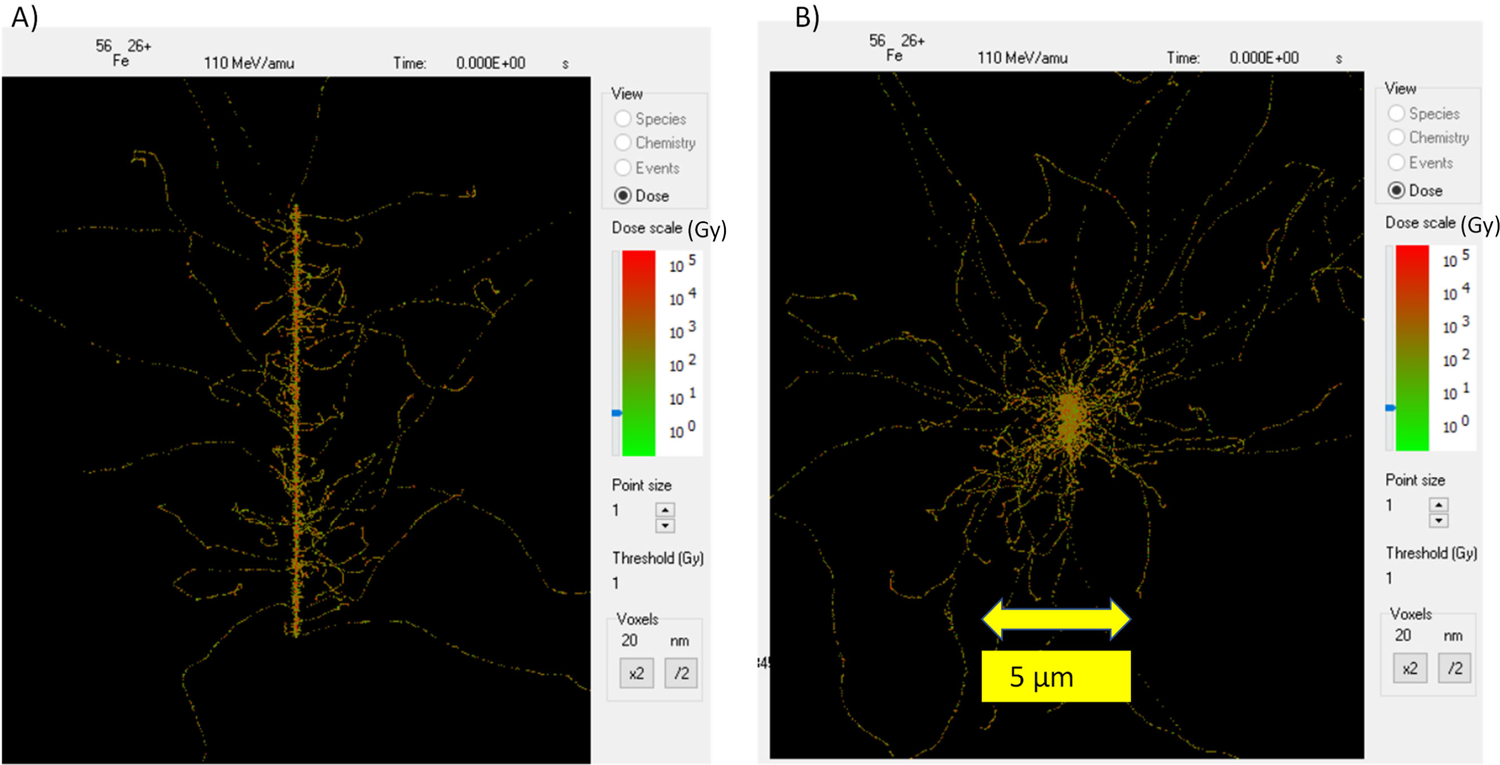
Representative track structure for ^56^Fe at LET=456 keV/μm from simulation of RITRACKS computer code [30]. Panel A) longitudinal view of 10 μm track length. Panel B) lateral view of identical track as Panel A).

Models for tissue specific ERR for cancer incidence are taking directly from the published reports from the Radiation Effects Research Foundation (RERF) for site-specific cancer incidence [50–59]. Statistical and dosimetry errors in the model parameters from these reports are used directly in our Monte-Carlo uncertainty analysis. Because additive risk models were not reported for several of the tissues in these reports [50–59] and to be consistent with the multiplicative model use for circulatory risk estimates [27] we report on the multiplicative model results in this paper.

A Bayesian approach is used to combine the DDREF estimate from the atomic-bomb survivor data with estimates from mouse tumor induction [18,20]. We studied the BEIR VII estimates and one by Hoel [60] and combined these with mouse data for solid cancers in mice and found small differences on their impact to GCR risk predictions due to the dominance of the core like terms [20]. For NSCR-2022 we use the Bayesian derived PDF with a mean DDREF=2.

Solid tumor data in mice are used directly to model the value and PDF for the parameter Ʃ_0_/α_γ_, where the PDF is represented by a three-parameter logistic function [20]. We use values derived excluding the Harderian Gland (HG) tumors as this organ does not appear in humans, and hepatocellular carcinoma in male mice because it was found to be an outlier from values for other cancers. HG tumor response data are used in estimation of other *R_QF_* parameters because it is the most detailed data set for radiation quality dependence for tumor induction [8,39]. The parameter values and uncertainties for the parameters *m* and *κ* are estimated from experiments on detailed radiation quality dependences with 5 ion’s or more in cell culture for chromosomal aberrations, neoplastic transformation, and HPRT mutations and the HG mouse tumor induction [20,21,38]. Using the constraint of Eq. (6) shows that the value of *m* has small influence on results for the range of 2 to 4. The value of the κ parameter is set at a higher value for light ions (κ=1000) based on experiments with fission neutrons and low energy light ions (<10 MeV/u). This difference in the κ parameter value between light and heavy ions is attributed to the influence of the varying effectiveness of δ-rays of different energies with these energies increasing with ion velocity. Electrons with energies below ∼20 keV are observed to have increased biological effectiveness [61,62], which plays a larger role for lower energy ions (<10 MeV/u) compared to relativistic energy ions. This energy region makes a large contribution for tracks of low energy light ions, which are abundant from slowing down of GCR primaries and secondaries from neutrons and other ions. In contrast, heavy ions have low abundance and short ranges below kinetic energies of 10 MeV/u and therefore are not a major contributor to space radiation risks.

### 7. Predictions for Space Missions

We considered astronauts of age 35-years at first missions and age 40-years at second mission. In a recent publication [23] it was shown that US Whites have the highest risks due to their higher background cancer rates and life-table, while US Blacks having a modestly lower radiation risk compared to US Whites. US Asian-Pacific Islanders (API) were found to be at the lowest radiation risks in the multiplicative model, with US Hispanics slightly higher. We therefore studied US Whites and US API groups as representative of the most and least sensitive model populations. GCR exposures are slowly varying for shielding below 100 g/cm^2^ of aluminum and we used 30 g/cm^2^ for an ISS exposure and 20 g/cm^2^ for lunar segments. We assumed 180-day ISS missions and considered GCR and trapped protons as described previously [13]. For possible lunar missions we used a slow Earth to moon transit time of 10 days and a 60-day surface time based on reported scenarios [63]. Average solar cycle conditions are used for ISS segment while an average solar minimum is used for the lunar segment.

**Table 2** shows predictions of REIC and **Table 3** for REID with the inclusion of circulatory disease risk predictions. Results predict NTE increase REIC by about ∼2.3 fold over the model that ignored NTEs. The predictions with NTEs show the lack of REID <3% with a 95% confidence interval for multiple ISS and lunar missions. Comparison of US Whites to US API show a decrease in cancer risks of about 30% compared to US whites. Cancer risks for multiple missions are slightly less than additive, which is due to declining risk with age at exposure and increased competition with background risks as radiation risks increase. Females have higher risks compared to males largely due to breast and ovarian cancers, and higher risk for lung cancer as predicted by low LET epidemiology data. For males the use of a higher RBE for male liver cancers based on mouse studies would narrow this difference to some extent as liver cancers makeup about ∼10% of the overall cancer risk. This difference in high LET biological effectiveness for liver cancer between males and females requires further study.

**TABLE 2.** Risk of Exposure Induced Cancer (REIC) estimates for average solar cycle on ISS and average solar min on lunar surface mission. Average shielding of 30 g/cm^2^ of aluminum for ISS and 20 g/cm^2^ of aluminum for lunar mission segments are used in calculations.

| **Mission** | **%REIC**<br>**without NTE** | **%REIC**<br>**with NTE** | **%REIC**<br>**without NTE** | **%REIC**<br>**with NTE** |
| --- | --- | --- | --- | --- |
|  | **Female (White)** |  | **Male (White)** |  |
| **ISS (180 d)** | 2.15 [0.64, 6.37] | 4.87 [0.26,11.2] | 1.21 [0.37,3.62] | 2.75 [0.96,6.1] |
| **Lunar (80 d)** | 1.7 [0.47, 5.26] | 3.33 [1.09,8.09] | 1.0 [0.29, 3.07] | 1.92 [0.67,4.56] |
| **ISS + Lunar** | 3.75 [1.05,11.5] | 5.87 [2.27,16.2] | 2.07 [0.64,6.1] | 3.84 [1.36,9.04] |
|  | **Female (Asian-Pacific Islanders)** |  | **Male (Asian-Pacific Islanders)** |  |
| **ISS (180 d)** | 1.68 [0.49,4.98] | 3.82 [1.26,8.83] | 0.91 [0.27, 2.73] | 2.1 [0.74,4.7] |
| **Lunar (80 d)** | 1.34 [0.36,4.14] | 2.63 [0.85,6.46] | 0.75 [0.21,2.29] | 1.47 [0.51,3.5] |
| **ISS + Lunar** | 2.82 [0.82,8.45] | 5.27 [1.76,12.6] | 1.55 [0.48,4.61] | 2.94 [1.05,6.93] |

**Table 3.** Risk of Exposure Induce Death (REID) estimates with TE or NTE+TE models. Average shielding of 30 g/cm^2^ of aluminum for ISS and 20 g/cm^2^ of aluminum for lunar mission segments are used in calculations.

| **Mission** | **%REID,**<br>**Cancer** | **%REID,**<br>**Circulatory**<br>**Disease** | **%REID,**<br>**Total** | **%REID,**<br>**Cancer** | **%REID,**<br>**Circulatory**<br>**Disease** | **%REID,**<br>**Total** |
| --- | --- | --- | --- | --- | --- | --- |
|  | **Female (White) without NTE** |  |  | **Male (White) without NTE** |  |  |
| **ISS (180 d)** | 0.68 [0.21,1.99] | 0.19 [0.07,0.37] | 0.88 [0.36,2.25] | 0.44 [0.14,1.3] | 0.23 [0.09,0.46] | 0.68 [0.31,1.6] |
| **Lunar (80 d)** | 0.54 [0.15,1.66] | 0.15 [0.06,0.28] | 0.70 [0.27,1.88] | 0.36 [0.1,1.1] | 0.19 [0.07,0.36] | 0.55 [0.24,1.34] |
| **ISS + Lunar** | 1.12 [0.36,3.46] | 0.33 [0.13,0.64] | 1.51 [0.62,3.87] | 0.75 [0.24,2.18] | 0.4 [0.16,0.78] | 1.16 [0.53,2.68] |
|  | **Female (Asian-Pacific Islanders) without NTE** |  |  | **Male (Asian-Pacific Islanders) without NTE** |  |  |
| **ISS (180 d)** | 0.48 [0.15,1.41] | 0.21 [0.08,0.42] | 0.70 [0.31, 1.7] | 0.33 [0.1,0.98] | 0.24 [0.09,0.46] | 0.57 [0.26,1.29] |
| **Lunar (80 d)** | 0.39 [0.11, 1.18] | 0.16 [0.06, 0.32] | 0.56 [0.23,1.43] | 0.27 [0.08,0.82] | 0.19 [0.07,0.37] | 0.47 [0.21,1.07] |
| **ISS+Lunar** | 0.83 [0.25,2.46] | 0.37 [0.15,0.73] | 1.12 [0.54,2.94] | 0.56 [0.17,1.65] | 0.41 [0.16, 0.8] | 0.98 [0.45,2.17] |
|  | **Female (White) with NTE** |  |  | **Male (White) with NTE** |  |  |
| **ISS (180 d)** | 1.53 [0.54,3.48] | 0.19 [0.07, 3.7] | 1.72 [0.7,3.67] | 0.98 [0.34,2.19] | 0.23 [0.09,0.46] | 1.22 [0.55,2.44] |
| **Lunar (80 d)** | 1.05 [0.36,2.53] | 0.14 [0.06,0.28] | 1.2 [0.49,2.67] | 0.69 [0.24,1.6] | 0.18 [0.07,0.35] | 0.87 [0.4,1.8] |
| **ISS+Lunar** | 1.9 [0.76,5.11] | 0.32 [0.12,0.63] | 2.5 [1.1,5.54] | 1.37 [0.49,3.24] | 0.4 [0.15,0.77] | 1.78 [0.82,3.65] |
|  | **Female (Asian-Pacific Islanders)**<br>**with NTE** |  |  | **Male (Asian-Pacific Islanders)**<br>**with NTE** |  |  |
| **ISS (180 d)** | 1.09 [0.38,2.46] | 0.21 [0.08,0.42] | 1.31 [0.57,2.7] | 0.75 [0.27,1.68] | 0.23 [0.09,0.46] | 0.99 [0.46,1.93] |
| **Lunar (80 d)** | 0.76 [0.26,1.84] | 0.16 [0.06,0.32] | 0.93 [0.4,2.02] | 0.52 [0.18,1.24] | 0.19 [0.07,0.37] | 0.71 [0.34,1.45] |
| **ISS + Lunar** | 1.54 [0.55,3.61] | 0.37 [0.14,0.73] | 1.92 [0.87,4.02] | 1.05 [0.38,2.47] | 0.41 [0.16,0.8] | 1.47 [0.7,2.92] |

The inclusion of circulatory risks increases mortality estimates about 25% and 37% for females and males, respectively in the model ignoring NTEs, and 20% and 30% when NTEs are assumed to modify RBE’s for solid cancers. Circulatory disease risks for US Whites and US API are predicted to be nearly the same. At this time RBE’s for circulatory diseases or their possible surrogate endpoints are extremely sparse, which leads to an uncertainty that is difficult to quantify. Using the NCRP recommended RBE model for non-cancer risks [11,29] suggests the proportion of circulatory disease risk in the overall risk is lower for space radiation compared to low LET exposures on Earth. A healthy worker effect for circulator risk found for astronauts would also suggest a lower proportion [15]. In addition, we note that differences in treatment of circulatory diseases with time period and exposed group leads to uncertainty when applying historical epidemiology data to astronauts in current or future missions, while the impact of such differences for cancer risk has been estimated using the BEIR VII [25] recommended approach shown in Eq. (3).

### 8. Summary

Cancer risk is the dominant risk established for long-term space missions. The predictions made here for combined ISS and lunar missions suggest risks are within recommendations of the NCRP for such missions [11,17] whose value for radiation protection of astronauts was described in a recent report [64]. Participation in a 3^rd^ long duration mission could possibly exceed the NCRP recommendations for ISS and lunar operations, however would be dependent on solar cycle, time between missions, age, sex, and possibly other factors. An important issue for cancer risk assessment is a possible deviation from a linear response for high LET radiation at low doses due to NTEs, which impacts RBE estimates. NTEs include bystander effects where cells traversed by heavy ions transmit oncogenic signals to nearby cells, genomic instability in the progeny of irradiated cells, and tissue microenvironment changes related to cancer development [5–10].

Mechanistic studies that have used low doses (<0.1 Gy), where less than one particle traverses a cell nucleus, suggests NTE dominate low dose risk from high LET radiation. A concern is the potential limitation of experimental models to quantify NTEs, however we note these are similar to the models used in assigning radiation quality factors in the past. The effects of NTEs are shown to significantly increase risk predictions for ISS and lunar missions in the present paper and for a Mars mission in a previous report [21]. A major concern is that most radiobiology studies continue to employ heavy ion doses corresponding to more than one particle per cell nucleus (>∼0.1 Gy) that will not be sensitive to NTEs, while GCR exposures occur where NTE’s are predicted to dominate risks. Organ absorbed doses from heavy ions are less than 0.1 Gy for a Mars mission, and less than 0.05 Gy for multiple ISS and lunar mission scenarios. Clearly the development of sensitive experimental models and data sets at low doses of heavy ions (<0.1 Gy) is needed to improve risk assessments. In addition, it is recommended the use of relativistic super heavy ions (A>58) such as high-energy Nb, La or Pb beams at low doses should be investigated as an optimal tool to investigate NTEs for space radiobiology applications.

## Data Availability

All data produced in the present work are contained in the manuscrip

## References

1. NCRP. Information needed to make radiation protection recommendations for space missions beyond Low-Earth orbit. National Council on Radiation Protection and Measurements Report No. 153: Bethesda MD, 2006.

2. Cucinotta FA, Durante M. Cancer risk from exposure to galactic cosmic rays: implications for space exploration by human beings. Lancet Oncol 2006;7:431–5.

3. Durante M, Cucinotta FA. Heavy ion carcinogenesis and human space exploration. Nature Rev Cancer 2008;8:465–72.

4. Kim MY, Hayat ML, Feiveson AH, Cucinotta FA. Using high-energy proton fluence to improve risk prediction for consequences of solar particle events. Adv Space Res 2009;44:1428–32.

5. Kadhim M, Salomaa S, Wright EG, Hildenbrandt G, Belyakov OV, et al., Non-targeted effects of ionizing radiation-implications for low dose risk. Mut Res 2013;752:84–98.

6. Lorimore SA, Coates PJ, Wright EG. Radiation-induced genomic instability and bystander effects: inter-related nontargeted effects of exposure to ionizing radiation. Oncogene 2003;22:7058–69.

7. Barcellos-Hoff MH, Mao J. HZE radiation non-targeted effects on the microenvironment that mediate mammary carcinogenesis. Front Oncol 2016; 6:57.

8. Chang PY, Cucinotta FA, Bjornstad KA, Bakke CJ, Du N, Fairchild DG, E. Cacao, et al. Harderian gland tumorigenesis: low-dose- and LET-Response. Radiat Res 2016;185:448–59.

9. Hada M, Chappell LJ, Wang M, George KA, Cucinotta FA. On the induction of chromosomal aberrations at fluence of less than one HZE particle per cell nucleus. Radiat Res 2014;182:68–379.

10. Sridharan DM, Asaithamby A, Bailey SM, Costes S, Doetsch PW, et al. Understanding cancer development processes following HZE particle exposure: roles of ROS, DNA damage repair, and inflammation. Radiat Res 2015; 183: 1–26.

11. NCRP. Recommendations of dose limits for low earth orbit. National Council on Radiation Protection and Measurements. NCRP Report 132: Bethesda MD, 2000.

12. Dietze G, Bartlett DT, Cool DA, Cucinotta FA, Jia X, McAulay IR, et al. Assessment of astronaut exposures in space. ICRP Publication 123. Ed. C. Clement. Annals of the ICRP 2013;: 42(4).

13. Cucinotta FA, Kim MH, Willingham V, George KA. Physical and biological organ dosimetry analysis for International Space Station astronauts. Radiat Res 2008;170:127–38.

14. NAS, Radiation Hazards to Crews on Crews on Interplanetary Missions, Natl Acad Press, Washington, D. C., 1996.

15. Cucinotta FA, Kim MY, Chappell L. Space radiation cancer risk projections and uncertainties-2012. NASA Technical Paper 2013-217375.

16. NRC, Technical evaluation of the NASA model for cancer risk to astronauts due to space radiation. National Research Council. The National Academies Press, Washington DC, 2013.

17. NCRP. Radiation protection for space activities: supplement to previous recommendations. National Council on Radiation Protection and Measurements Commentary 23: Bethesda MD, 2014.

18. Cucinotta FA. A new approach to reduce uncertainties in space radiation cancer risk predictions. PLoS One 2015;10(3): e0120717.

19. Cucinotta FA. Space radiation risks for astronauts on multiple International Space Station missions. PLoS One 2014;9(4): e96099.

20. Cucinotta FA, To K, Cacao E. Predictions of space radiation fatality risk for exploration missions. Life Sci Space Res 2017;13:1–11.

21. Cucinotta FA, Cacao E. Non-targeted effects models predict significantly higher Mars mission cancer risk than targeted effects models. Sci Rep 2017; 7:1832.

22. Cucinotta FA, Cacao E, Kim MY, Saganti PB. Benchmarking risk predictions and uncertainties in the NSCR model of GCR cancer risks with revised low LET risk coefficients. Life Sci Space Res 2020;27:64–73.

23. Cucinotta FA, Saganti PB. Race and ethnic group dependent space radiation cancer risk predictions. Scientific Rep 2023;12:2028.

24. Cucinotta FA. Flying without a net: space radiation cancer risk predictions without a gamma-ray basis. Int J Mol Sci 2022;23(8):4324.

25. BEIR VII. Health risks from exposure to low levels of ionizing radiation. National Academy of Sciences Committee on the Biological Effects of Radiation. Washington DC: National Academy of Sciences Press; 2006.

26. Cucinotta FA, Kim MY, Chappell LJ, Huff JL. How safe is safe enough? radiation protection for a human mission to Mars. PLoS One 2013;8(10): e74988.

27. Little MP, Azizova TV, Richardson DB, Tapio S, Bernier M, Kreuzer M, et al. Ionising radiation and cardiovascular disease: systematic review and meta-analysis. British Med J 2023;380:e072924.

28. Little M, Azizova D, Bazyka S, Bouffler SD, Cardis E, et al. Meta-analysis of circulatory disease from exposure to low-level ionizing radiation and estimates of potential population risks. Environ Health Persp 2012;120:1503–11.

29. NCRP. Operational Radiation Safety Program for Astronauts in Low-Earth. NCRP Report 142, Bethesda MD, 2002.

30. Plante I, Cucinotta FA. Ionization and excitation cross sections for the interaction of HZE particles in liquid water and application to Monte-Carlo simulation of radiation tracks. New J Phys 2008;10:1–15.

31. Katz R, Ackerson B, Homayoonfar M, Scharma SC. Inactivation of cells by heavy ion bombardment. Radiat Res 1971;47:402–25.

32. Katz R. High LET constraints on low LET survival. Phys Med Biol 1978;23:909–16.

33. Cucinotta FA, Nikjoo H, Goodhead DT. Comment on the effects of delta-rays on the number of particle-track transversals per cell in laboratory and space exposures. Radiat Res 1998;150:115–9.

34. NIST. Stopping-power and range tables for electrons, protons, and helium ions. National Institute of Standards and Technology Report NISTIR 4999; Gaithersburg MD, 2009.

35. Barkas H. Nuclear Research Emulsions. Academic Press Inc, New York 1963; Vol. 1, Chap. 9, p,371.

[36. Goodhead DT. Relationship of radiation track structure to biological effect: a re-interpretation of the parameters of the Katz model. Nuclear Tracks Radiat Meas 1989;116:177–84.

37. Cucinotta FA. Biophysics of NASA Radiation Quality Factors. Radiat Protect Dosim 2015;166(1-4):282–9.

38. Cacao E, Hada M, Saganti PB, George KA, Cucinotta FA. Relative biological effectiveness of HZE particles for chromosomal exchanges and other surrogate cancer risk endpoints. PLoS One 2016;11(4): e0153998.

39. Alpen EL, Powers-Risius P, Curtis SB, DeGuzman R. Tumorigenic potential of high-Z, high-LET charged particle radiations. Radiat Res 1993;88:132–43.

40. Bettega D, Calzoolari P, NorisChiorda G., 1992. Tallone-Lombardi. Transformation of C3H10T1/2 cells with 4.3 MeV α particles at low doses: effects of single and fractionated doses. Radiat. Res. 1992;131:66–71.

41. Nagasawa H, Little JB. Bystander effect for chromosomal aberrations induced in wild-type and repair deficient CHO cells by low fluences of alpha particles. Mutat Res 2002;508:121–9.

42. Belyakov OV, Mitchell SA, Parikh D, et al. Biological effects in unirradiated human tissue induced by radiation damage up to 1mm away. Proc Natl Acad Sci USA. 2005;102:14203–8.

43. Tsuboi K, Yang TC, Chen DJ. Charged-particle mutagenesis I. Cytoxi and mutagenic effects of high-LET charged particles on human skin fibroblasts. Radiat Res 1992;129:171–176.

44. Cucinotta FA, Katz R, Wilson JW, Dubey RD. Radial dose distributions in the delta-ray theory of track structure In: Proceedings of Two Center Effects in Ion-Atom Collisions. AIP Conference Proceedings, AIP Press. Ed. by T. J. Gay and A.F. Starace, 245–265, 1996.

45. Varma MN, Baum JW. Energy deposition in nanometer eegions by 377 MeV/nucleon neon ions. Radiat Res 1980;81:355–363.

46. Wilson JW, Townsend LW, Shinn JL, Cucinotta FA, Costen RC, et al. Galactic cosmic ray transport methods, past, present, and future. Adv Space Res 1994; 10:841–52.

47. Cucinotta FA, Kim MY, Schneider SI, Hassler DM. Description of light ion production cross sections and fluxes on the Mars surface using the QMSFRG model. Radiat Environ Biophys 2007;46:101–6.

48. Zeitlin C, Hassler DM, Cucinotta FA, Ehresmann B, Wimmer-Schweingruber RF, et al. Measurements of the energetic particle radiation environment in transit to Mars on the Mars Science laboratory. Science 2013;340:1080–84.

49. Kim MY, Cucinotta FA, Nounu H, Zeitlin C, Hassler DM, et al. Comparison of Martian surface ionizing radiation measurements from MSL-RAD with Badhwar-O’Neill 2011/HZETRN model calculations. J Geophys Res 2014;119:1311–21

50. Hsu W, Preston DL, Soda M, Sugiyama H, Funamoto S, et al., The incidence of leukemia, lymphoma, and multiple myeloma among atomic bomb survivors: 1950-2001. Radiat Res 2013;179:361–82.

51. Grant EJ, Brenner A, Sugiyama H, Sakata R, Sadakane A, Utada M, et al. Solid cancer incidence among the Life-span study of atomic-bomb survivors: 1958-2009. Radiat Res 2017;187:513–537.

52. Cahoon EK, Preston DL, Pierce DA, Grant E, Brenner AV, et al. Lung, laryngeal and other respiratory cancer incidence among Japanese atomic bomb survivors: an updated analysis from 1958 through 2009. Radiat Res 2017;187:538–48.

53. Brenner AV, Preston DL, Sakata R, Sugiyama H, Berrington de Gonzalez A, et al. Incidence of breast cancer in the Life Span Study of the atomic bomb survivors: 1958-2009. Radiat Res 2018;190:433–444.

54. Sadakane A, French B, Brenner AV, Preston DL, Sugiyama H, et al,. Radiation and risk of liver, biliary tracts, and pancreatic cancers in the atomic bomb survivors in Hiroshima and Nagasaki: 1958-2009. Radiat Res 2019;192:299–310.

55. Sakata R, et al. Radiation-related risk of cancers of the upper digestive tract among Japanese atomic bomb survivors. Radiat Res 2019;192:331–344.

56. Brenner AV, Sugiyama H, Preston DL, Sakata R, French B, et al. Radiation risk of central nervous system tumors in the Life Space Study of atomic bomb survivors, 1958-2009. Eur J Epidem 2020;35:591-600.

57. Sugiyami H, Misumi M, Brenner A, Grant EJ, Sakata R, et al. Radiation risk of incident colorectal cancer by anatomic site among atomic bomb survivors: 1958-2009. Int J Cancer 2019;146:635–45.

58. Mabuchi K, Preston DL, Brenner AV, Sugiyama H, Utada M, et al. Risk of prostate cancer incidence among atomic bomb survivors: 1958-2009. Radiat Res 2021;195:66–76.

59. Utada M, Brenner AV, Preston DL, Cologne JB, Sakata R, et al. Radiation risk of ovarian cancer in atomic bomb survivors: 1958-2009. Radiat Res 2021;195:60–5.

60. Hoel DG. Comments on the DDREF estimate of the BEIR VII committee. Health Phys 25;108:351–6.

61. Nikjoo H, Goodhead DT. Track structure analysis illustrating the prominent role of low energy electrons in radiobiological effects of low-LET radiations. Phys Med Biol 1991;36:229–38.

62. Goodhead DT. Biological effectiveness of lower-energy photons for cancer risk. Radiat Prot Dosim 2019;183:197–202.

63. Lunar missions web site: https://www.nasa.gov/specials/artemis/

64. Cucinotta FA, Schimmerling W, Blakely EA, Hei T. A proposed change to astronaut exposure limits is a giant leap back for radiation protection. Life Sci Space Res 2021;31:59–70.

